# High-throughput multivariable Mendelian randomization analysis prioritizes apolipoprotein B as key lipid risk factor for coronary artery disease

**DOI:** 10.1101/2020.02.10.20021691

**Authors:** Verena Zuber, Dipender Gill, Mika Ala-Korpela, Claudia Langenberg, Adam Butterworth, Leonardo Bottolo, Stephen Burgess

## Abstract

**Background:** Genetic variants can be used to prioritize risk factors as potential therapeutic targets via Mendelian randomization (MR). An agnostic statistical framework using Bayesian model averaging (MR-BMA) can disentangle the causal role of correlated risk factors with shared genetic predictors. Here, our objective is to identify lipoprotein measures as mediators between lipid-associated genetic variants and coronary artery disease (CAD) for the purpose of detecting therapeutic targets for CAD.

**Methods:** As risk factors we consider 30 lipoprotein measures and metabolites derived from a high-throughput metabolomics study including 24,925 participants. We fit multivariable MR models of genetic associations with CAD estimated in 453,595 participants (including 113,937 cases) regressed on genetic associations with the risk factors. MR-BMA assigns to each combination of risk factors a model score quantifying how well the genetic associations with CAD are explained. Risk factors are ranked by their marginal score and selected using false discovery rate (FDR) criteria. We perform supplementary and sensitivity analyses varying the dataset for genetic associations with CAD.

**Results:** In the main analysis, the top combination of risk factors ranked by the model score contains apolipoprotein B (ApoB) only. ApoB is also the highest ranked risk factor with respect to the marginal score (FDR< 0.005). Additionally, ApoB is selected in all sensitivity analyses. No other measure of cholesterol or triglyceride is consistently selected otherwise.

**Conclusions:** Our agnostic genetic investigation prioritizes ApoB across all datasets considered, suggesting that ApoB, representing the total number of hepatic-derived lipoprotein particles, is the primary lipid determinant of CAD.

**Key messages:**

- It is a common consensus that certain lipoproteins increase cardiovascular disease risk, yet the exact mechanisms are unclear.
- We use genetic associations with high-throughput metabolomics features to draw a detailed picture of lipid traits and characteristics allowing for an unprecedented resolution when considering lipids as risk factors for cardiovascular disease.
- This study integrates genetic data from a large scale metabolomics study including 25,000 samples and the largest study on cardiovascular disease risk including 113,937 cases and 339,658 controls.
- Mendelian randomization – Bayesian model averaging (MR-BMA), a novel algorithm for multivariable Mendelian randomization is used to identify the most likely causal lipid determinants of cardiovascular disease from a large set of candidate risk factors with shared genetic predictors.
- Our agnostic genetic investigation prioritizes apolipoprotein B across all datasets considered, suggesting that apolipoprotein B, representing the total number of hepatic-derived lipoprotein particles, is the primary lipid determinant of cardiovascular disease risk.

## Introduction

Genetic variants have the potential to contribute greatly to our understanding of mechanisms underlying disease processes, and to guide target validation for pharmacological and clinical interventions that reduce disease risk [1]. Coronary artery disease (CAD) is the most common cause of death globally. While it has been shown that genetic variants predisposing individuals to higher levels of low-density lipoproteins (LDL)-cholesterol also associate with increased CAD risk [2], genetic variants predisposing individuals to higher levels of high-density lipoproteins (HDL)-cholesterol are not associated with CAD risk [3] after accounting for other lipid traits. These genetic analyses may suggest that LDL-cholesterol is a causal risk factor for CAD risk, but HDL-cholesterol is not – as has generally been observed in clinical trials of lipid-altering therapies [4, 5, 6]. Genetic studies have also suggested that triglyceride levels are an independent risk factor for CAD risk [7]. Triglycerides are another component of body fat which are transported by lipoprotein particles, and in particular by very low density lipoproteins (VLDL). However, two recent studies showed that genetic associations with CAD risk are proportional to the change in apolipoprotein B (ApoB), the primary protein component of VLDL, LDL, and intermediate-density lipoprotein (IDL) particles, and that LDL-cholesterol and triglycerides do not appear to be independent risk factors for CAD after accounting for ApoB [8, 9].

Genome-wide association studies (GWAS) are increasingly used to combine genomic profiling with high-throughput molecular measures on a large scale, including tens of thousands of samples, to explore the genetic regulation of molecular processes. For example, Kettunen et al. have combined high-throughput metabolomics with genomic profiling on nearly 25 000 individuals [10]. Given the large sample size, these studies are well powered to explore the causal role of molecular mechanisms. The metabolomics study by Kettunen et al. was conducted using nuclear magnetic resonance (NMR) spectroscopy to provide a detailed characterization of lipid-related traits, including 14 size categories of lipoprotein particles ranging from small HDL to extra-extra-large VLDL. For each lipoprotein category, measurements are available of cholesterol, triglycerides, cholesterol ester, and phospholipid content. Additional mean diameter of the lipoprotein particles is measured for some lipoprotein size categories. Measurements also include apolipoprotein A1 and ApoB, sphingomyelins, fatty acids, and phosphoglycerides (Supplementary Table S1).

Previous MR studies on lipid determinants for CAD risk have included only a few curated lipid traits at a time [8, 9]. In this study, we build on a high-throughput metabolomics data resource [10] to investigate a much wider set of lipoprotein measurements as candidate risk factors for CAD. We use a recently published algorithm called Mendelian randomization with Bayesian model averaging (MR-BMA) [11] that applies principles from high-dimensional data analysis and machine learning to detect causal risk factors from a large set of candidate risk factors. Our goal is to select the lipoprotein measures that are the most likely causal risk factors for CAD.

## Methods

### Variable selection method for finding likely causal risk factors

We provide a brief outline of the MR-BMA method here. More details are given in the Supplementary Material, and a diagram illustrating the method is shown in Figure 1.

**Figure 1:**
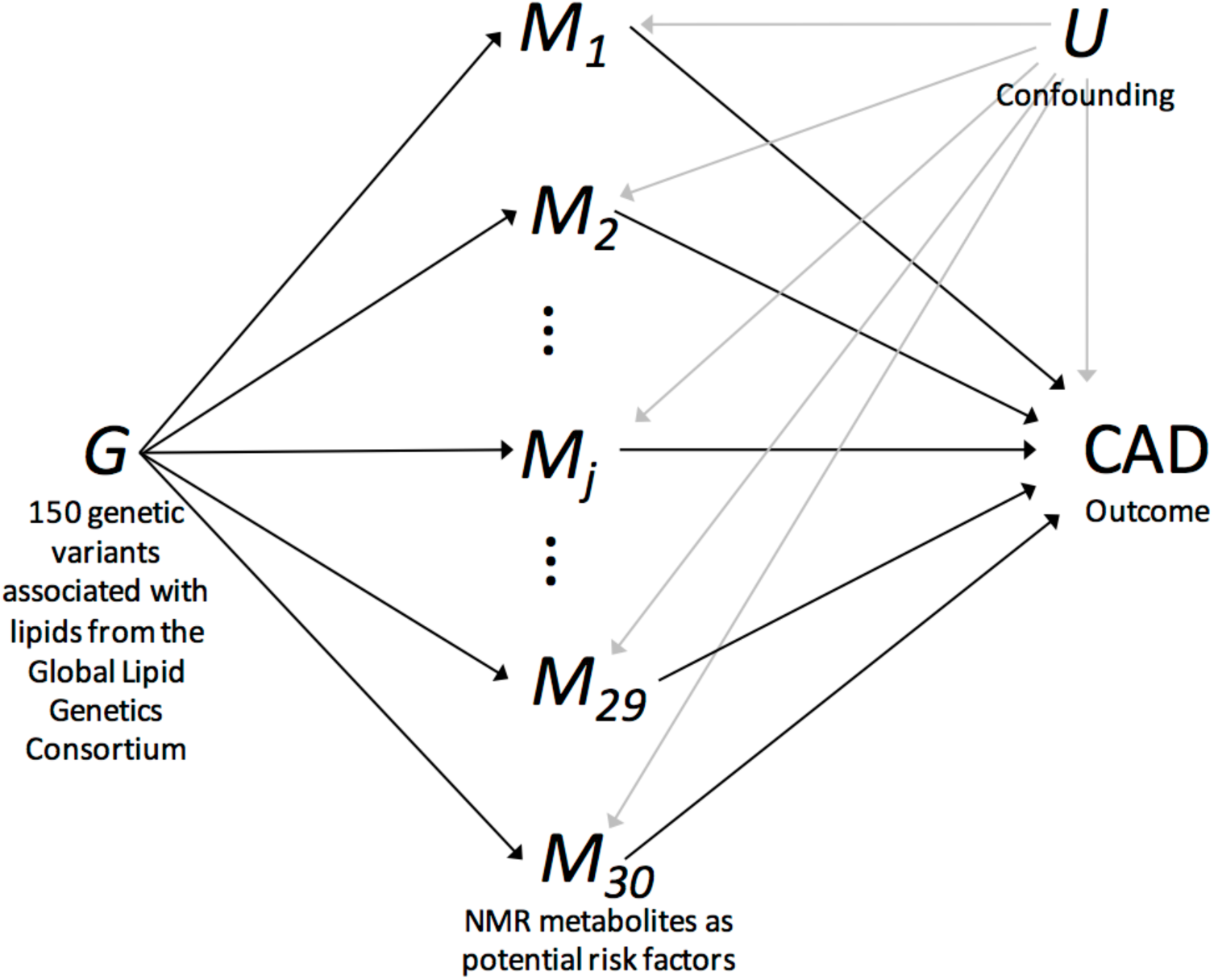
Diagram illustrating multivariable Mendelian randomization for selecting causal risk factors for coronary artery disease (CAD) from a large number of candidate risk factors, e.g. metabolites measured using nuclear magnetic resonance (NMR) spectroscopy. Legend: *G* = genetic variants, *M*_1_, …, *M*_30_ = metabolites as risk factors, *CAD* = coronary artery disease as outcome, *U* = confounders.

We consider each set of risk factors in turn: all single risk factors, all pairs of risk factors, all triples, and so on. For each set of risk factors, we undertake a multivariable MR analysis using weighted regression based on summarized genetic data. We assess goodness-of-fit in the regression model, and assign a score to the risk factor set that is the model posterior probability of that set being the true causal risk factors. We repeat this to get a posterior probability for all models (i.e. all sets of risk factors). Then, for each of the candidate risk factors, we sum up the posterior probability over models including that risk factor to compute the marginal inclusion probability for the risk factor, representing the probability of that risk factor being a causal determinant of disease risk. We also calculate the model-averaged causal effect, representing the average causal effect across models including that risk factor. P-values are calculated for each risk factor using a permutation method, with adjustment for multiple testing via the Benjamini and Hochberg false discovery rate (FDR) procedure [12].

### Study design

A summary of our study design is given in Figure 2. The three key steps in designing a two-sample multivariable MR study are instrument selection, risk factor selection, and the choice of outcome data, including main and sensitivity analysis.

**Figure 2:**
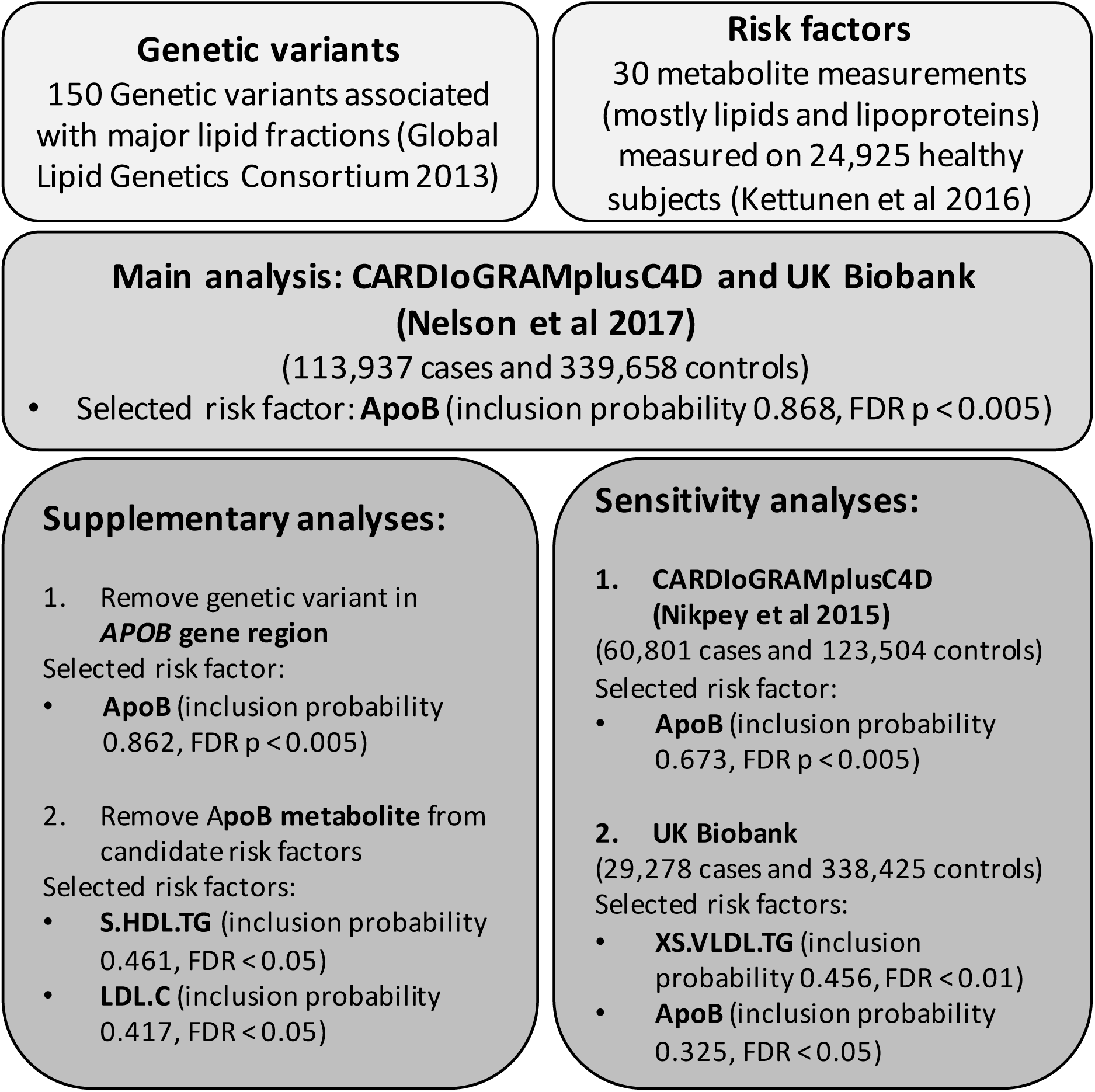
Schematic diagram of the study design and results for the main, supplementary, and sensitivity analyses. Selected risk factors are those which had a empirical *p*-value of less than 0.05 after correction for multiple testing.

### Selecting lipid-associated variants as instrumental variables

We took an initial list of 185 variants associated with blood lipids (LDL-cholesterol, HDL-cholesterol or triglycerides) in the Global Lipid Genetics Consortium at a genome-wide level of significance (*p* < 5 × 10^−8^) [7] which was pruned at a linkage disequilibrium threshold of *r*^2^ < 0.05, and further refined by genomic distance, excluding variants that are less than 1 megabase pair apart, to provide a list of *n* = 150 genetic variants. We selected these lipid-associated genetic variants as instrumental variables because we wanted to investigate lipid determinants of CAD risk. This is important to keep in mind when interpreting the results as the prioritization of risk factors by MR-BMA is conditional on the genetic variants selected as instrumental variables. There are two direct consequences of this choice. Firstly, this choice of instrumental variables will downweight non-lipid risk factors, and so results should not be interpreted as evidence that those risk factors are not on the causal path to CAD. Secondly, basing the selection of instrumental variables on an external dataset (e.g. the Global Lipid Genetics Consortium) reduces the risk of winner’s course [13].

We performed supplementary analysis with *n* = 55 genetic variants derived from the NMR GWAS as instrumental variables to investigate how much the results depend on the choice of instruments.

### Lipoprotein measures as risk factors

Genetic associations with lipoprotein measures and metabolites are taken from Kettunen et al. [10] who measured 118 variables on 24,925 European individuals using the high-throughput Nightingale NMR platform. The contributing cohorts are mainly Finish (around 50%), with several other Belgium (10%), Dutch (16%), Estonian (14%) and German (10%) cohorts contributing. The majority of samples for measuring the biomarkers were collected after overnight-fasting, otherwise the analyses were adjusted for time to last meal [10]. Estimates were obtained by linear regression of each NMR measurement on each of the genetic variants in turn, with adjustment for age, sex, time from last meal (if non-fasting), and ten genomic principal components. NMR measurements were inverse rank-based normal transformed, so that association estimates are presented in standard deviation units for the relevant risk factor throughout.

Several measurements from the Nightingale platform were highly correlated, judged by the correlation between the genetic associations for the 150 genetic variants. While MR-BMA was able to identify the causal risk factors reliably in a simulation study when risk factors were highly correlated (up to |*r*| = 0.99) [11], several risk factors were more highly correlated than this. We therefore pruned the set of risk factors to avoid inaccurate results due to collinearity. For each lipoprotein diameter category representing the size of lipoproteins, we retained only the measurement of cholesterol and/or triglyceride content, and mean particle diameter where available. We also included only total fatty acid content and not other fatty acid measurements, as genetic predictors that were able to distinguish reliably between these risk factors were not included as instruments. Other non-lipoprotein metabolite measurements were retained in the analysis as they had substantially weaker correlations with lipoprotein measurements, and so would only be selected by MR-BMA if they mediated CAD risk from the genetic predictors included in the model. No pair of risk factors included in the final analysis were more highly correlated than |*r*| = 0.99 (see correlation heatmap in Supplementary Figure S1). Finally, we only included risk factors into the MR analysis that had at least one genetic variant that was a strong predictor (genome-wide significant). The final list of 30 lipoprotein measures and metabolites included in the analysis is provided in Supplementary Table S1.

### Coronary artery disease as outcome

Our primary analysis was based on genetic associations with CAD risk taken from the 2017 CARDIoGRAMplusC4D data release meta-analysed together with UK Biobank [14] including 453,595 individuals mostly of European descent, of whom 113,937 had a CAD event. Genetic association estimates with CAD risk were obtained in each study of the CARDIo-GRAMplusC4D consortium by logistic regression with adjustment for at least five genomic principal components, and then meta-analysed across studies. There was one rare genetic variant (rs1998013, effect allele frequency 0.8%, in the *PCSK9* gene region) and one common intergenic genetic variant (rs894210, effect allele frequency 43.5%) for which there was no association estimate with CAD available. After excluding the missing genetic variants, we performed MR-BMA with 148 variants and 30 risk factors.

As supplementary analyses, we repeated the same analysis steps on the 2017 CARDIo-GRAMplusC4D data release except: 1) we omitted the variant in the *APOB* gene region from the analysis, to assess whether this variant was overly influential in determining the top ranked models and 2) we omitted the ApoB measurement from the list of risk factors to see if any other risk factor reached a similar level of evidential support. If it is the case that ApoB was selected as representative for a group of highly correlated traits, then upon removal of ApoB another risk factor of this group should be selected as representative instead.

As sensitivity analyses, we considered 1) an earlier release of CARDIoGRAMplusC4D consortium [15] including 60,801 CAD cases and 123,504 controls of European descent, but not including UK Biobank participants and 2) a UK Biobank GWAS which includes 29,278 cases and 338,425 controls of European descent (defined by self-report and genomic principal components). Quality control procedures were performed and related individuals were excluded from the analysis as described previously [16]. For the main and the two sensitivity analyses we report the results including all variants and after excluding genetic variants that are influential points and outliers.

An overview of the data sources used to derive genetic associations with CAD risk is provided in Supplementary Table S2.

## Results

### Main analysis using outcome data from CARDIoGRAMplusC4D and UK Biobank

Results are provided in Table 1. We show the top 10 models (i.e. sets of risk factors) ranked according to their model posterior probability, and the top 10 risk factors according to their marginal inclusion probability. We also present the model-averaged causal effect estimate for each risk factor. The top-ranked model contains ApoB and no additional risk factors (model posterior probability 0.464). ApoB is also the risk factor with the strongest overall evidence (marginal inclusion probability 0.868, FDR< 0.005). A diagnostic scatterplot of the genetic associations with the outcome against the genetic associations with ApoB is given in Figure 3. Our primary analysis was performed after model diagnostics, which removed influential (Supplementary Table S3 and Supplementary Figure S2) and outlying genetic variants (Supplementary Table S4 and Supplementary Figure S3) from the analysis. Similar results were obtained including all variants in the analysis (Supplementary Table S5).

**Table 1:** Main analysis: Top 10 models (combination of risk factors) ranked by the model posterior probability and top 10 risk factors ranked by the marginal inclusion probability in the primary analysis based on *n* = 138 genetic variants after model diagnostics. Causal effects are log odds ratios for coronary artery disease per 1 standard deviation increase in the risk factor. Empirical *p*-values are computed using 1, 000 permutations and adjusted for multiple testing using False Discovery Rate (FDR) procedure.

| CARDIoGRAMplusC4D and UK Biobank |  |  |  |  |  |  |  |  |
| --- | --- | --- | --- | --- | --- | --- | --- | --- |
| | Model | Posterior probability | Causal effect | Risk factor | Marginal inclusion probability | Model-averaged causal effect | Empirical $p$-value | FDR |
| 1 | ApoB | 0.480 | 0.464 | ApoB | 0.868 | 0.392 | 0.0001 | 0.003 |
| 2 | ApoB,S.HDL.TG | 0.043 | 0.349,0.175 | S.HDL.TG | 0.136 | 0.033 | 0.0165 | 0.247 |
| 3 | LDL.C,S.HDL.TG | 0.021 | 0.276,0.301 | LDL.C | 0.075 | 0.015 | 0.0882 | 0.882 |
| 4 | ApoB,M.HDL.C | 0.020 | 0.437,-0.111 | XXL.VLDL.TG | 0.047 | 0.010 | 0.4823 | 0.995 |
| 5 | ApoB,S.LDL.C | 0.014 | 0.570,-0.121 | Serum.C | 0.045 | 0.011 | 0.2295 | 0.995 |
| 6 | ApoB,XXL.VLDL.TG | 0.014 | 0.419,0.112 | IDL.C | 0.042 | 0.008 | 0.2401 | 0.995 |
| 7 | ApoB,XS.VLDL.TG | 0.011 | 0.375,0.099 | S.LDL.C | 0.040 | 0.001 | 0.3745 | 0.995 |
| 8 | ApoB,S.VLDL.C | 0.011 | 0.480,-0.017 | M.HDL.C | 0.038 | -0.005 | 0.2885 | 0.995 |
| 9 | ApoB,LDL.C | 0.011 | 0.522,-0.062 | HDL.C | 0.036 | -0.006 | 0.2266 | 0.995 |
| 10 | ApoB,HDL.C | 0.011 | 0.453,-0.073 | Serum.TG | 0.035 | 0.006 | 0.7583 | 0.995 |

**Figure 3:**
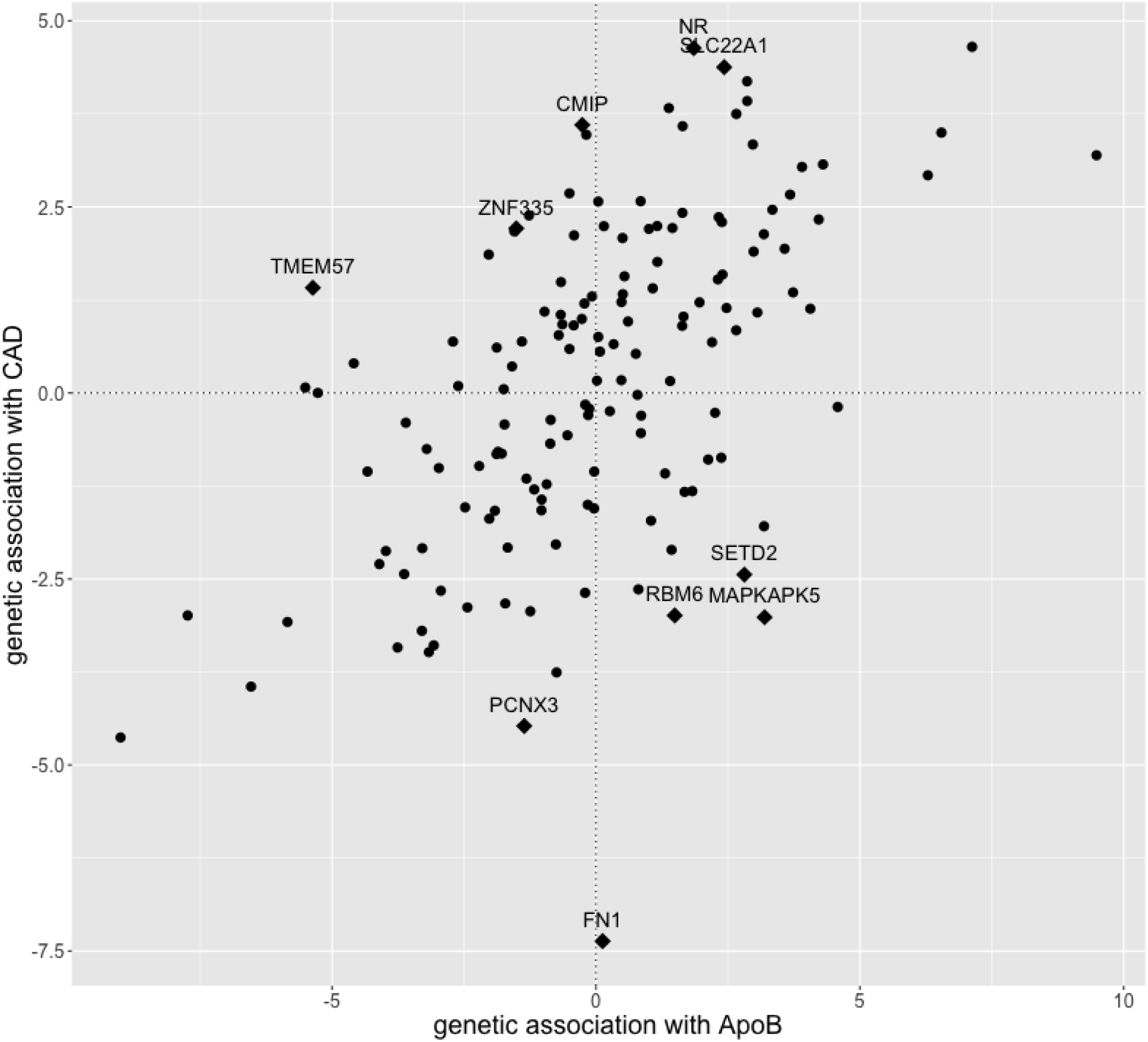
Estimates of genetic associations with coronary artery disease (CAD) risk (*y*-axis) against genetic associations with apolipoprotein B (ApoB, *x*-axis) for each genetic variant from the primary analysis using CARDIoGRAMplusC4D and UK Biobank. Outliers removed from the analysis are highlighted as diamonds (♦) and their annotated gene-region is displayed.

### Supplementary analysis

As supplementary analyses, we first repeated the primary analysis excluding the genetic variant in the *APOB* gene region, to ensure that this variant was not driving the selection of ApoB as a risk factor. This exclusion did not impact the results (Supplementary Table S6) – ApoB remained the highest ranking individual model (model posterior probability 0.455) and the risk factor with the strongest marginal evidence (marginal inclusion probability 0.862). Secondly, we repeated the primary analysis excluding ApoB from the list of risk factors (Supplementary Table S7). No alternative risk factor had similar strength of evidence, suggesting that ApoB is indeed the most important risk factor and not just a representative of a group of highly correlated lipoprotein measures with similar evidence. On exclusion of ApoB, the top risk factors were triglycerides content in small HDL particles (marginal inclusion probability 0.461, FDR< 0.05) and LDL cholesterol (marginal inclusion probability 0.417, FDR< 0.05). Yet, the evidence for these two lipoprotein measures is much weaker compared to the evidence for ApoB in the main analysis.

As final supplementary analysis we used *n* = 55 genetic variants derived from the NMR GWAS as instrumental variables. After removing influential variants and outlying genetic variants, ApoB is the risk factor with the strongest evidence (Supplementary Table S8). This supplementary analysis has less power than the main analysis which is due to a smaller number of genetic variants used as instrumental variables (*n* = 55 based on NMR GWAS while *n* = 148 based on Global Lipids Genetics Consortium), yet it confirms ApoB as the highest ranked risk factor. It is important to consider here the different interpretation which depends on the selection of instruments. For the main analysis instrumental variables were selected based on the Global Lipids Genetics Consortium with the aim to define lipid-related risk factors for CAD. The supplementary analysis in contrast allows for a wider panel of NMR metabolites as risk factors for CAD.

### Sensitivity analysis

As sensitivity analysis, we used genetic associations with CAD risk from two alternative datasets. Results are shown in Supplementary Table S9. For the earlier release of CARDIo-GRAMplusC4D [15], the top ranked model includes ApoB alone (model posterior probability 0.455), and ApoB is the top ranked risk factor overall (marginal inclusion probability 0.673, FDR< 0.005). For UK Biobank, ApoB (marginal inclusion probability 0.325, FDR< 0.05) was ranked second after triglycerides in very small VLDL-cholesterol (marginal inclusion probability 0.456, FDR< 0.01). When looking at the individual models, triglycerides content in very small VLDL-cholesterol particles is ranked first followed by models including both ApoB and a measure of triglycerides content, suggesting an additional causal pathway via triglycerides when deriving genetic associations from UK Biobank analysis.

## Discussion

Our results add to the growing evidence that ApoB is the primary causal determinant for CAD risk amongst lipoprotein measurements [17, 8, 18]. Cholesterol underlies the development of atherosclerosis [19]. It enters the arterial wall within those ApoB-containing lipoprotein particles that are small enough to enter the tunica intima from the circulation; these particles include small VLDL, IDL and LDL particles as well as lipoprotein(a). The recent genetic evidence, together with the results in this work, strongly point towards the direction that the lipid content of the particles in secondary to ApoB [8, 18, 20].

These results do not invalidate LDL-cholesterol as a causal risk factor for CAD risk. Indeed, LDL particles contain an apolipoprotein B molecule, as do IDL and VLDL particles. ApoB (in particular ApoB-100) represents the total number of hepatic-derived lipoprotein particles [21]. However, this investigation suggests clinical benefit of lowering triglyceride and LDL-C levels is proportional to the absolute change in ApoB. ApoB measurements are independent of particle density, and are not affected by heterogeneity of particle cholesterol content [22]. This is particularly important for accurately capturing the number of small dense LDL particles, which are believed to be associated with atherosclerosis. ApoB has been shown to be a superior measure to LDL-cholesterol in the prediction of CAD risk [23], and in prediction of coronary artery calcification [24]. From a clinical perspective, statins target LDL-cholesterol levels rather than ApoB, suggesting that greater benefit might be obtained from lipid-lowering drugs that target lipoprotein particle number [25]. When analysing data from UK Biobank only, there was also some evidence for triglyceride content measures as an additional risk factor. This was not evident in the main analysis or the sensitivity analysis including data from the earlier CARDIoGRAMplusC4D release. This finding should therefore be interpreted with some caution.

There are some caveats to the interpretation of the results of this study. Although we were able to distinguish between measures of cholesterol content and triglyceride content for some categories of lipoprotein particles, we were not able to distinguish between other lipoprotein measures, such as cholesterol ester and phospholipid content, which correlated almost perfectly with cholesterol content. Previous studies have suggested that the ApoB/ApoA1 ratio may be a relevant risk factor for CAD [26]. However, working on summary-level data we were not able to investigate any other relevant risk factors than those provided by the original data, such as the ratio ApoB/ApoA1 or the ratio of LDL/HDL particles. A further limitation is that there is overlap between individuals in the outcome datasets for the main and sensitivity analyses. For this reason, we refer to them as sensitivity analyses rather than replication analyses, as they assess the robustness of the variable selection algorithm to variations in the outcome dataset, rather than providing an independent replication of the findings.

One key strength of the study is that the genetic associations were derived on mainly fasting samples, which facilitates the interpretation of the results. Fasting measurements represent hepatic derived lipid traits while non-fasting samples also partly reflect gut-derived contributions from chylomicron particles and their remnants. This means that our analysis is well placed to answer causal questions about endogenous lipid pathways, but is less able to answer questions about lipoproteins from dietary sources.

In conclusion, our agnostic investigation to identify risk factors for CAD strongly prior-itized ApoB, suggesting that ApoB, representing the number of hepatic-derived lipoprotein particles, is the key determinant of CAD risk amongst lipid-related measurements. This analysis demonstrates the potential of publicly-available genetic association data from highthroughput experiments combined with modern data-adaptive statistical learning techniques for obtaining biological insights into disease aetiology.

## Data Availability

This study is based on publicly available data.

## Acknowledgment

This work was supported by the UK Medical Research Council (MC UU 00002/7). S.B. and V.Z. are supported by Sir Henry Dale Fellowship jointly funded by the Wellcome Trust and the Royal Society (Grant Number 204623/Z/16/Z).

D.G. was supported by the Wellcome Trust 4i Programme (203928/Z/16/Z) and British Heart Foundation Centre of Research Excellence (RE/18/4/34215) at Imperial College London. M.A.K was supported by a research grant from the Sigrid Juselius Foundation, Finland. This work was supported by core funding from: the UK Medical Research Council (MR/L003120/1), the British Heart Foundation (RG/13/13/30194; RG/18/13/33946), the National Institute for Health Research [Cambridge Biomedical Research Centre at the Cambridge University Hospitals NHS Foundation Trust] ^⋆^ and Health Data Research UK, which is funded by the UK Medical Research Council, Engineering and Physical Sciences Research Council, Economic and Social Research Council, Department of Health and Social Care (England), Chief Scientist Office of the Scottish Government Health and Social Care Directorates, Health and Social Care Research and Development Division (Welsh Government), Public Health Agency (Northern Ireland), British Heart Foundation and Wellcome.

^⋆^ The views expressed are those of the authors and not necessarily those of the NHS, the NIHR or the Department of Health and Social Care.

L.B. was supported by the BHF-Turing Cardiovascular Data Science Awards 2017 and The Alan Turing Institute under the Engineering and Physical Sciences Research Council grant EP/N510129/1.

The research has been conducted using the UK Biobank Resource under Application Number 26865. Lastly, this study would not have been possible without the access to publicly available summary data. We would like to thank the authors of the NMR GWAS (http://www.computationalmedicine.fi/data) and the CARDIOGRAMplus4CD (http://www.cardiogramplusc4d.org/data-downloads/).

