## Supplemental Data 1 for "High-throughput multivariable Mendelian randomization analysis prioritizes apolipoprotein B as key lipid risk factor for coronary artery disease"

### List of Tables

S1 List of lipoprotein and metabolite measurements included in the analyses. . . . S2
S2 Study summary table . . . S3
S3 Cook's distance to detect influential genetic variants . . . S4
S4 $q$-statistic to detect outlying genetic variants . . . S5
S5 Analysis including all genetic variants . . . S6
S6 Supplementary analysis 1 . . . S7
S7 Supplementary analysis 2 . . . S8
S8 Supplementary analysis 3 . . . S9
S9 Sensitivity analysis . . . S10
S10 Parameter check: Prior variance . . . S11
S11 Parameter check: Prior probability . . . S12

### List of Figures

S1 Genetic correlation between lipoprotein measures and metabolites . . . S13
S2 Diagnostic plots with Cooks distance . . . S14
S3 Diagnostic plots with $q$-statistic . . . S15

### Supplementary Tables

| Abbreviation | Lipoprotein and metabolite measurements included |
|---|---|
| XXL.VLDL.TG | Triglyceride content in chylomicrons and extra-extra large VLDL |
| XL.VLDL.TG | Triglyceride content in extra-large VLDL |
| L.VLDL.TG | Triglyceride content in large VLDL |
| M.VLDL.TG | Triglyceride content in medium VLDL |
| S.VLDL.TG | Triglyceride content in small VLDL |
| XS.VLDL.TG | Triglyceride content in extra-small VLDL |
| IDL.TG | Triglyceride content in IDL |
| XL.HDL.TG | Triglyceride content in extra-large HDL |
| S.HDL.TG | Triglyceride content in small HDL |
| Serum.TG | Serum total triglycerides |
| L.VLDL.C | Cholesterol content in large VLDL |
| M.VLDL.C | Cholesterol content in medium VLDL |
| S.VLDL.C | Cholesterol content in small VLDL |
| LDL.C | Cholesterol content in LDL |
| S.LDL.C | Cholesterol content in small LDL |
| IDL.C | Cholesterol content in IDL |
| XL.HDL.C | Cholesterol content in extra-large HDL |
| L.HDL.C | Cholesterol content in large HDL |
| M.HDL.C | Cholesterol content in medium HDL |
| HDL.C | Cholesterol content in HDL |
| Est.C | Esterified cholesterol |
| Serum.C | Serum total cholesterol |
| VLDL.D | VLDL diameter |
| LDL.D | LDL diameter |
| HDL.D | HDL diameter |
| ApoA1 | Apolipoprotein A1 |
| ApoB | Apolipoprotein B |
| SM | Sphingomyelins |
| Tot.FA | Total fatty acids |
| Tot.PG | Total phosphoglycerides |

Table S1: List of lipoprotein and metabolite measurements included in the analyses.

| | First author | Year | Pubmed | $n$ | Cases | Controls | Study names (population) |
|---|---|---|---|---|---|---|---|
| Risk factor | | | | | | | |
| | Kettunen | 2016 | 27005778 | 24,925 | | | NMR GWAS (European, predominantly Finnish) |
| Outcome | | | | | | | |
| Main | Nelson | 2017 | 28714975 | 453,595 | 113,937 | 339,658 | CARDIoGRAMplusC4D (European, South&East Asian) and UK Biobank (European) |
| Sensitivity | Nikpay | 2015 | 26343387 | 184,305 | 60,801 | 123,504 | CARDIoGRAMplusC4D (European, South&East Asian) |
| | UK Biobank (European) | 2019 | 31756303 | 367,703 | 29,278 | 338,425 | UK Biobank (European) |

Table S2: Study summary table

CARDIoGRAMplusC4D and UK Biobank

| | rs | gene region | $Cd1$ | $Cd2$ | $Cd3$ | max $Cd$ |
|---|---|---|---|---|---|---|
| 1 | rs10903129 | TMEM57 | 0.108 | 0.054 | 0.018 | 0.108 |
| 2 | rs2923084 | AMPD3 | 0.049 | 0.069 | 0.068 | 0.069 |
| 3 | rs6489818 | MAPKAPK5 | 0.051 | 0.029 | 0.004 | 0.051 |
| 4 | rs1515110 | NR | 0.013 | 0.042 | 0.027 | 0.042 |
| 5 | rs515135 | APOB | 0.013 | 0.003 | 0.041 | 0.041 |
| 6 | rs6859 | APOE | 0.035 | 0.018 | 0.039 | 0.039 |
| 7 | rs2326077 | intergenic | 0.039 | 0.027 | 0.015 | 0.039 |
| 8 | rs5880 | CETP | 0.001 | 0.038 | 0.023 | 0.038 |
| 9 | rs799160 | intergenic | 0.004 | 0.002 | 0.037 | 0.037 |
| 10 | rs4465830 | ZNF335 | 0.005 | 0.037 | 0.037 | 0.037 |
| | | threshold | 0.457 | 0.696 | 0.457 | |

CARDIoGRAMplusC4D only

| | rs | region | $Cd1$ | $Cd2$ | $Cd3$ | max $Cd$ |
|---|---|---|---|---|---|---|
| 1 | rs261342 | LIPC | 0.008 | 0.024 | 0.911 | **0.911** |
| 2 | rs5880 | CETP | 0.006 | 0.164 | 0.057 | 0.164 |
| 3 | rs515135 | APOB | 0.116 | 0.129 | 0.125 | 0.129 |
| 4 | rs2923084 | AMPD3 | 0.081 | 0.109 | 0.096 | 0.109 |
| 5 | rs10903129 | TMEM57 | 0.078 | 0.012 | 0.025 | 0.078 |
| 6 | rs4530754 | CSNK1G3 | 0.076 | 0.001 | 0.001 | 0.076 |
| 7 | rs6489818 | MAPKAPK5 | 0.062 | 0.005 | 0.009 | 0.062 |
| 8 | rs2326077 | intergenic | 0.039 | 0.016 | 0.015 | 0.039 |
| 9 | rs12133576 | DR1 | 0.036 | 0.006 | 0.004 | 0.036 |
| 10 | rs4465830 | ZNF335 | 0.005 | 0.036 | 0.000 | 0.036 |
| | | threshold | 0.457 | 0.457 | 0.457 | |

UK Biobank only

| | rs | region | $Cd1$ | $Cd2$ | $Cd3$ | $Cd4$ | max $Cd$ |
|---|---|---|---|---|---|---|---|
| 1 | rs10401969 | SUGP1 | 0.302 | 0.224 | 0.248 | 0.096 | 0.302 |
| 2 | rs2923084 | AMPD3 | 0.124 | 0.064 | 0.026 | 0.079 | 0.124 |
| 3 | rs5880 | CETP | 0.107 | 0.033 | 0.025 | 0 | 0.107 |
| 4 | rs2297374 | SLC22A1 | 0.024 | 0.054 | 0.091 | 0.057 | 0.091 |
| 5 | rs10903129 | TMEM57 | 0.012 | 0.051 | 0.005 | 0.071 | 0.071 |
| 6 | rs7703051 | HMGCR | 0.006 | 0.053 | 0.009 | 0.065 | 0.065 |
| 7 | rs6489818 | MAPKAPK5 | 0.005 | 0.032 | 0.001 | 0.055 | 0.055 |
| 8 | rs894210 | intergenic | 0.05 | 0.051 | 0.019 | 0.039 | 0.051 |
| 9 | rs687339 | intergenic | 0.038 | 0.044 | 0.039 | 0.045 | 0.045 |
| 10 | rs998584 | VEGFA | 0.041 | 0.039 | 0.037 | 0.036 | 0.041 |
| | | threshold | 0.457 | 0.457 | 0.696 | 0.457 | |

Table S3: Influential genetic variants: This table displays for each study the 10 variants with the largest Cook's distance ($Cd$) and the annotated genomic region based on the best individual models (model posterior probability > 0.02). The maximum $Cd$ of each variant in all models is used for diagnostics. The final row gives the suggested cut-off for Cook's distance and genetic variants with $Cd$ above the threshold are marked in bold.

CARDIoGRAMplusC4D and UK Biobank

| | rs | gene region | $q1$ | $q2$ | $q3$ | max $q$ |
|---|---|---|---|---|---|---|
| 1 | rs1250229 | FN1 | 55.077 | 54.867 | 57.211 | **57.211** |
| 2 | rs6489818 | MAPKAPK5 | 19.308 | 20.150 | 12.288 | **20.150** |
| 3 | rs12801636 | PCNX3 | 15.124 | 14.625 | 15.845 | **15.845** |
| 4 | rs1515110 | NR | 14.697 | 10.106 | 11.196 | **14.697** |
| 5 | rs2290547 | SETD2 | 13.361 | 14.316 | 8.53 | **14.316** |
| 6 | rs2297374 | SLC22A1 | 11.075 | 11.676 | 14.204 | **14.204** |
| 7 | rs10903129 | TMEM57 | 13.910 | 12.503 | 7.369 | **13.910** |
| 8 | rs2925979 | CMIP | 13.787 | 11.646 | 10.338 | **13.787** |
| 9 | rs2240327 | RBM6 | 13.213 | 11.505 | 11.223 | **13.213** |
| 10 | rs4465830 | ZNF335 | 8.194 | 2.964 | 12.962 | **12.962** |
| 11 | rs6450176 | ARL15 | 8.271 | 6.936 | 12.705 | 12.705 |
| 12 | rs731839 | PEPD | 12.596 | 10.504 | 10.14 | 12.596 |
| 13 | rs4148218 | ABCG8 | 11.789 | 12.03 | 12.032 | 12.032 |
| 14 | rs2247056 | HLA | 8.710 | 9.897 | 11.563 | 11.563 |
| 15 | rs9930333 | FTO | 7.213 | 6.599 | 11.191 | 11.191 |
| | | threshold | | | | 12.84801 |

CARDIoGRAMplusC4D only

| | rs | gene region | $q1$ | $q2$ | $q3$ | max $q$ |
|---|---|---|---|---|---|---|
| 1 | rs4530754 | CSNK1G3 | 24.505 | 15.468 | 15.292 | **24.505** |
| 2 | rs6489818 | MAPKAPK5 | 19.513 | 14.598 | 13.255 | **19.513** |
| 3 | rs12801636 | PCNX3 | 16.290 | 16.800 | 16.810 | **16.810** |
| 4 | rs4148218 | ABCG8 | 14.936 | 14.107 | 15.098 | **15.098** |
| 5 | rs1250229 | FN1 | 9.932 | 12.776 | 10.769 | 12.776 |
| 6 | rs952044 | AC090771.2 | 10.333 | 12.468 | 11.714 | 12.468 |
| 7 | rs2297374 | SLC22A1 | 9.125 | 9.187 | 11.492 | 11.492 |
| 8 | rs4465830 | ZNF335 | 7.196 | 5.401 | 11.390 | 11.390 |
| 9 | rs998584 | VEGFA | 8.781 | 11.195 | 7.745 | 11.195 |
| 10 | rs2923084 | AMPD3 | 8.404 | 10.802 | 9.845 | 10.802 |
| | | threshold | | | | 12.84801 |

UK Biobank only

| | rs | gene region | $q1$ | $q2$ | $q3$ | $q4$ | max $q$ |
|---|---|---|---|---|---|---|---|
| 1 | rs2297374 | SLC22A1 | 38.863 | 34.587 | 27.820 | 34.345 | **38.863** |
| 2 | rs1250229 | FN1 | 25.528 | 22.807 | 31.310 | 24.057 | **31.310** |
| 3 | rs6489818 | MAPKAPK5 | 14.222 | 18.563 | 12.616 | 20.625 | **20.625** |
| 4 | rs2240327 | RBM6 | 15.063 | 16.844 | 16.278 | 17.034 | **17.034** |
| 5 | rs687339 | intergenic | 16.284 | 15.452 | 7.003 | 15.328 | **16.284** |
| 6 | rs2925979 | CMIP | 10.424 | 15.160 | 9.803 | 13.903 | **15.160** |
| 7 | rs4148218 | ABCG8 | 14.250 | 14.512 | 11.681 | 14.137 | **14.512** |
| 8 | rs4921914 | NAT2 | 13.259 | 10.640 | 10.262 | 11.642 | **13.259** |
| 9 | rs1186380 | HNF1A-AS1 | 9.758 | 12.067 | 11.982 | 13.168 | **13.168** |
| 10 | rs2241210 | UBE3B | 12.630 | 11.045 | 10.759 | 9.015 | 12.630 |
| | | threshold | | | | | 12.87313 |

Table S4: Outlying genetic variants: This table displays for each study the 10 variants with the largest maximum $q$ and the annotated genomic region based on the best individual models (model posterior probability > 0.02). The maximum $q$ of each variant in all models is used for diagnostics. The final row gives the suggested threshold for the $q-$statistic and variants with $q$ above this threshold are given in bold.

| CARDIoGRAMplusC4D and UK Biobank | | | | | | |
|---|---|---|---|---|---|---|
| | Model | Posterior probability | Causal effect | Risk factor | Marginal inclusion probability | Model-averaged causal effect |
| 1 | ApoB | 0.347 | 0.432 | ApoB | 0.706 | 0.298 |
| 2 | ApoB,M.HDL.C | 0.048 | 0.392,-0.17 | M.HDL.C | 0.124 | -0.024 |
| 3 | XS.VLDL.TG | 0.039 | 0.411 | XS.VLDL.TG | 0.103 | 0.032 |
| 4 | ApoB,S.LDL.C | 0.015 | 0.613,-0.208 | IDL.TG | 0.079 | 0.021 |
| 5 | ApoB,SM | 0.014 | 0.501,-0.139 | XXL.VLDL.TG | 0.076 | 0.02 |
| 6 | IDL.TG | 0.014 | 0.38 | IDL.C | 0.074 | 0.018 |
| 7 | ApoB,S.HDL.TG | 0.014 | 0.334,0.151 | LDL.C | 0.052 | 0.005 |
| 8 | ApoB,XS.VLDL.TG | 0.014 | 0.287,0.163 | Serum.TG | 0.049 | 0.014 |
| 9 | ApoB,XXL.VLDL.TG | 0.013 | 0.37,0.156 | Serum.C | 0.048 | 0.009 |

| CARDIoGRAMplusC4D only | | | | | | |
|---|---|---|---|---|---|---|
| | Model | Posterior probability | Causal effect | Risk factor | Marginal inclusion probability | Model-averaged causal effect |
| 1 | ApoB | 0.24 | 0.438 | ApoB | 0.488 | 0.197 |
| 2 | XS.VLDL.TG | 0.058 | 0.42 | IDL.TG | 0.159 | 0.048 |
| 3 | IDL.TG | 0.033 | 0.395 | XS.VLDL.TG | 0.153 | 0.05 |
| 4 | S.VLDL.C | 0.015 | 0.447 | Serum.TG | 0.095 | 0.036 |
| 5 | ApoB,XS.VLDL.TG | 0.014 | 0.272,0.186 | Tot.FA | 0.088 | 0.026 |
| 6 | ApoB,S.HDL.TG | 0.012 | 0.331,0.163 | IDL.C | 0.076 | 0.016 |
| 7 | ApoB,IDL.TG | 0.012 | 0.283,0.167 | S.HDL.TG | 0.07 | 0.016 |
| 8 | IDL.TG,XXL.VLDL. | 0.012 | 0.319,0.256 | XXL.VLDL.TG | 0.067 | 0.016 |
| 9 | ApoB,M.HDL.C | 0.01 | 0.407,-0.127 | Serum.C | 0.065 | 0.016 |
| 10 | ApoB,Serum.TG | 0.01 | 0.318,0.16 | S.LDL.C | 0.064 | 0.011 |

| UK Biobank only | | | | | | |
|---|---|---|---|---|---|---|
| | Model | Posterior probability | Causal effect | Risk factor | Marginal inclusion probability | Model-averaged causal effect |
| 1 | XS.VLDL.TG | 0.205 | 0.459 | XS.VLDL.TG | 0.388 | 0.161 |
| 2 | S.VLDL.C | 0.032 | 0.488 | Tot.FA | 0.321 | 0.139 |
| 3 | HDL.C,Tot.FA | 0.03 | -0.255,0.475 | ApoB | 0.147 | 0.047 |
| 4 | ApoB | 0.023 | 0.452 | IDL.TG | 0.145 | 0.045 |
| 5 | IDL.TG | 0.019 | 0.425 | HDL.C | 0.103 | -0.023 |
| 6 | ApoB,XS.VLDL.TG | 0.014 | 0.191,0.294 | S.VLDL.C | 0.099 | 0.033 |
| 7 | L.HDL.C,Tot.FA | 0.013 | -0.221,0.448 | S.HDL.TG | 0.097 | 0.026 |
| 8 | S.HDL.TG,Tot.FA | 0.011 | 0.329,0.259 | TotPG | 0.089 | -0.032 |
| 9 | Tot.FA,TotPG | 0.01 | 0.883,-0.504 | IDL.C | 0.073 | 0.015 |
| 10 | LDL.C,XS.VLDL.TG | 0.009 | 0.129,0.369 | Serum.TG | 0.072 | 0.026 |

Table S5: Analysis including all genetic variants: Top 10 models ranked by the model posterior probability and top 10 risk factors ranked by the marginal inclusion probability including all genetic variants before removing influential genetic variants and outliers. Causal effects are log odds ratios for coronary artery disease per 1 standard deviation increase in the risk factor.

| | CARDIoGRAMplusC4D and UK Biobank | | | | | |
|---|---|---|---|---|---|---|
| | Model | Posterior probability | Causal effect | Risk factor | Marginal inclusion probability | Model-averaged causal effect |
| 1 | ApoB | 0.472 | 0.46 | ApoB | 0.862 | 0.385 |
| 2 | ApoB,S.HDL.TG | 0.043 | 0.343,0.177 | S.HDL.TG | 0.136 | 0.033 |
| 3 | LDL.C,S.HDL.TG | 0.02 | 0.272,0.301 | LDL.C | 0.076 | 0.015 |
| 4 | ApoB,M.HDL.C | 0.019 | 0.435,-0.11 | XXL.VLDL.TG | 0.05 | 0.011 |
| 5 | ApoB,XXL.VLDL.TG | 0.015 | 0.408,0.123 | Serum.C | 0.045 | 0.01 |
| 6 | ApoB,S.LDL.C | 0.015 | 0.571,-0.127 | IDL.C | 0.043 | 0.008 |
| 7 | ApoB,XS.VLDL.TG | 0.012 | 0.367,0.102 | S.LDL.C | 0.041 | 0.001 |
| 8 | ApoB,Serum.TG | 0.011 | 0.385,0.098 | Serum.TG | 0.038 | 0.007 |
| 9 | ApoB,LDL.C | 0.011 | 0.525,-0.071 | M.HDL.C | 0.037 | -0.004 |
| 10 | ApoB,S.VLDL.C | 0.011 | 0.474,-0.015 | HDL.C | 0.035 | -0.005 |

Table S6: Supplementary analysis 1: After excluding the genetic variant in the *APOB* gene region, these are the top 10 models judged by posterior probability and top 10 risk factors judged by marginal inclusion probability in the primary analysis based on $n = 137$ genetic variants. Causal effects are log odds ratios for coronary artery disease per 1 standard deviation increase in the risk factor.

| CARDIoGRAMplusC4D and UK Biobank including all genetic variants ($n$ = 148) | | | | | | |
|---|---|---|---|---|---|---|
| | Model | Posterior probability | Causal effect | Risk factor | Marginal inclusion probability | Model-averaged causal effect |
| 1 | XS.VLDL.TG | 0.127 | 0.411 | XS.VLDL.TG | 0.263 | 0.094 |
| 2 | IDL.TG | 0.046 | 0.38 | IDL.C | 0.213 | 0.059 |
| 3 | S.VLDL.C | 0.041 | 0.44 | IDL.TG | 0.204 | 0.063 |
| 4 | IDL.C,XXL.VLDL.TG | 0.03 | 0.299,0.347 | XXL.VLDL.TG | 0.168 | 0.05 |
| 5 | IDL.TG,XXL.VLDL.TG | 0.022 | 0.304,0.267 | M.HDL.C | 0.162 | -0.037 |
| 6 | M.HDL.C,Serum.C | 0.015 | -0.317,0.367 | Serum.C | 0.116 | 0.034 |
| 7 | LDL.C,XS.VLDL.TG | 0.015 | 0.178,0.286 | LDL.C | 0.114 | 0.026 |
| 8 | IDL.C,S.HDL.TG | 0.012 | 0.241,0.282 | Serum.TG | 0.107 | 0.038 |
| 9 | IDL.C,Serum.TG | 0.011 | 0.219,0.287 | S.VLDL.C | 0.096 | 0.031 |
| 10 | S.LDL.C,XS.VLDL.TG | 0.011 | 0.175,0.294 | S.HDL.TG | 0.079 | 0.019 |

| CARDIoGRAMplusC4D and UK Biobank after model diagnostics ($n$ = 138) | | | | | | | | |
|---|---|---|---|---|---|---|---|---|
| | Model | Posterior probability | Causal effect | Risk factor | Marginal inclusion probability | Model-averaged causal effect | Empirical $p-$value | FDR |
| 1 | LDL.C,S.HDL.TG | 0.156 | 0.261,0.3 | S.HDL.TG | 0.461 | 0.144 | 0.0021 | 0.025 |
| 2 | IDL.TG | 0.063 | 0.436 | LDL.C | 0.417 | 0.119 | 0.0013 | 0.025 |
| 3 | S.HDL.TG,S.LDL.C | 0.049 | 0.294,0.266 | Serum.C | 0.17 | 0.056 | 0.0143 | 0.087 |
| 4 | IDL.C,S.HDL.TG | 0.048 | 0.237,0.325 | IDL.TG | 0.159 | 0.055 | 0.0151 | 0.087 |
| 5 | L.HDL.C,Serum.C | 0.032 | -0.272,0.381 | S.LDL.C | 0.156 | 0.038 | 0.0274 | 0.101 |
| 6 | HDL.C,Serum.C | 0.027 | -0.277,0.441 | IDL.C | 0.128 | 0.029 | 0.0296 | 0.101 |
| 7 | S.HDL.TG,Serum.C | 0.021 | 0.354,0.23 | L.HDL.C | 0.118 | -0.026 | 0.0181 | 0.087 |
| 8 | LDL.C,XS.VLDL.TG | 0.016 | 0.233,0.249 | HDL.C | 0.095 | -0.019 | 0.0384 | 0.115 |
| 9 | Est.C,S.HDL.TG | 0.014 | 0.197,0.393 | XS.VLDL.TG | 0.076 | 0.017 | 0.0682 | 0.182 |
| 10 | LDL.C,XXL.VLDL.TG | 0.012 | 0.337,0.273 | XXL.VLDL.TG | 0.073 | 0.016 | 0.2636 | 0.575 |

Table S7: Supplementary analysis 2: After excluding the ApoB measurement as risk factor from the set of candidate risk factors these are the top 10 models ranked by the posterior probability and top 10 risk factors ranked by the marginal inclusion probability in the primary analysis based on all available genetic variants ($n$ = 148) and after model diagnostics ($n$ = 138). Causal effects are log odds ratios for coronary artery disease per 1 standard deviation increase in the risk factor.

| CARDIoGRAMplusC4D and UK Biobank including all NMR GWAS genetic variants ($n$ = 55) | | | | | | |
|---|---|---|---|---|---|---|
| | Model | Posterior probability | Causal effect | Risk factor | Marginal inclusion probability | Model-averaged causal effect |
| 1 | M.VLDL.C,XXL.VLDL.TG | 0.063 | 0.957,-1.018 | M.VLDL.C | 0.325 | 0.312 |
| 2 | IDL.TG | 0.061 | 0.397 | XXL.VLDL.TG | 0.32 | -0.267 |
| 3 | Serum.TG,XXL.VLDL.TG | 0.046 | 0.974,-1.025 | Serum.TG | 0.214 | 0.19 |
| 4 | L.VLDL.C,M.VLDL.C | 0.038 | -1.113,1.224 | XL.VLDL.TG | 0.173 | -0.13 |
| 5 | M.VLDL.C,XL.VLDL.TG | 0.036 | 0.966,-0.961 | IDL.TG | 0.149 | 0.057 |
| 6 | IDL.C | 0.036 | 0.348 | L.VLDL.C | 0.134 | -0.107 |
| 7 | ApoB | 0.023 | 0.369 | L.VLDL.TG | 0.115 | -0.083 |
| 8 | Serum.TG,XL.VLDL.TG | 0.02 | 0.974,-0.954 | IDL.C | 0.1 | 0.033 |
| 9 | L.VLDL.TG,Serum.TG | 0.016 | -0.998,1.065 | S.VLDL.C | 0.095 | 0.044 |
| 10 | S.VLDL.C,XXL.VLDL.TG | 0.013 | 0.565,-0.385 | XS.VLDL.TG | 0.068 | 0.029 |

| CARDIoGRAMplusC4D and UK Biobank after model diagnostics ($n$ = 45) | | | | | | | | |
|---|---|---|---|---|---|---|---|---|
| | Model | Posterior probability | Causal effect | Risk factor | Marginal inclusion probability | Model-averaged causal effect | Empirical $p-$value | FDR |
| 1 | ApoB | 0.155 | 0.3 | ApoB | 0.492 | 0.153 | 1.0E-04 | 0.004 |
| 2 | S.VLDL.C | 0.077 | 0.269 | S.VLDL.C | 0.246 | 0.068 | 4.0E-04 | 0.009 |
| 3 | ApoB,Crea | 0.047 | 0.318,-0.196 | Crea | 0.241 | -0.049 | 0.026 | 0.247 |
| 4 | Crea,S.VLDL.C | 0.025 | -0.201,0.287 | M.VLDL.C | 0.07 | 0.022 | 0.013 | 0.201 |
| 5 | XS.VLDL.TG | 0.015 | 0.227 | S.LDL.C | 0.069 | 0.014 | 0.030 | 0.247 |
| 6 | ApoB,Gly | 0.014 | 0.303,-0.064 | Phe | 0.067 | -0.011 | 0.137 | 0.441 |
| 7 | ApoB,Phe | 0.011 | 0.306,-0.157 | Gly | 0.067 | -0.004 | 0.043 | 0.276 |
| 8 | S.LDL.C | 0.01 | 0.329 | XS.VLDL.TG | 0.062 | 0.01 | 0.033 | 0.247 |
| 9 | M.VLDL.C | 0.008 | 0.225 | S.HDL.TG | 0.06 | 0.011 | 0.072 | 0.404 |
| 10 | S.HDL.TG | 0.007 | 0.244 | M.HDL.C | 0.06 | -0.01 | 0.117 | 0.441 |

Table S8: Supplementary analysis 3: We additionally varied our set of instruments and performed an analysis based on genetic variants associated with any of the measures in the metabolomics genome-wide association study by Kettunen et al. These are the top 10 models ranked by the posterior probability and top 10 risk factors ranked by the marginal inclusion probability in the primary analysis based on all available genetic variants ($n$ = 55) and after model diagnostics ($n$ = 45). Causal effects are log odds ratios for coronary artery disease per 1 standard deviation increase in the risk factor.

**CARDIoGRAMplusC4D only**

| | Model | Posterior probability | Causal effect | Risk factor | Marginal inclusion probability | Model-averaged causal effect | Empirical $p-$value | FDR |
|---|---|---|---|---|---|---|---|---|
| 1 | ApoB | 0.394 | 0.455 | ApoB | 0.673 | 0.293 | 0.0001 | 0.003 |
| 2 | ApoB,M.HDL.C | 0.018 | 0.425,-0.121 | LDL.C | 0.107 | 0.027 | 0.0544 | 0.461 |
| 3 | S.VLDL.C | 0.018 | 0.464 | S.LDL.C | 0.097 | 0.027 | 0.0709 | 0.461 |
| 4 | IDL.TG | 0.014 | 0.444 | Serum.TG | 0.084 | 0.028 | 0.0599 | 0.461 |
| 5 | HDL.C,Serum.C | 0.014 | -0.263,0.464 | Serum.C | 0.072 | 0.021 | 0.1176 | 0.510 |
| 6 | LDL.C,Serum.TG | 0.012 | 0.276,0.263 | HDL.C | 0.062 | -0.012 | 0.0974 | 0.506 |
| 7 | ApoB,Serum.TG | 0.011 | 0.369,0.115 | S.VLDL.C | 0.059 | 0.015 | 0.1667 | 0.542 |
| 8 | ApoB,IDL.TG | 0.011 | 0.358,0.109 | IDL.TG | 0.056 | 0.015 | 0.1539 | 0.542 |
| 9 | S.LDL.C | 0.010 | 0.461 | M.HDL.C | 0.055 | -0.008 | 0.1889 | 0.546 |
| 10 | ApoB,S.VLDL.C | 0.010 | 0.402,0.06 | IDL.C | 0.052 | 0.010 | 0.2423 | 0.630 |

**UK Biobank only**

| | Model | Posterior probability | Causal effect | Risk factor | Marginal inclusion probability | Model-averaged causal effect | Empirical $p-$value | FDR |
|---|---|---|---|---|---|---|---|---|
| 1 | XS.VLDL.TG | 0.195 | 0.435 | XS.VLDL.TG | 0.456 | 0.169 | 0.0002 | 0.006 |
| 2 | ApoB,S.HDL.TG | 0.056 | 0.281,0.233 | ApoB | 0.325 | 0.102 | 0.0010 | 0.015 |
| 3 | ApoB,XS.VLDL.TG | 0.043 | 0.207,0.258 | S.HDL.TG | 0.222 | 0.060 | 0.0061 | 0.061 |
| 4 | ApoB | 0.039 | 0.437 | IDL.TG | 0.109 | 0.027 | 0.0157 | 0.103 |
| 5 | LDL.C,XS.VLDL.TG | 0.024 | 0.14,0.338 | LDL.C | 0.108 | 0.018 | 0.0446 | 0.191 |
| 6 | S.VLDL.C | 0.024 | 0.467 | Serum.TG | 0.104 | 0.032 | 0.0171 | 0.103 |
| 7 | LDL.C,S.HDL.TG | 0.015 | 0.216,0.334 | S.VLDL.C | 0.086 | 0.024 | 0.0444 | 0.191 |
| 8 | S.LDL.C,XS.VLDL.TG | 0.015 | 0.133,0.346 | Tot.FA | 0.079 | 0.018 | 0.0677 | 0.254 |
| 9 | IDL.C,S.HDL.TG | 0.012 | 0.201,0.345 | IDL.C | 0.063 | 0.009 | 0.0994 | 0.331 |
| 10 | ApoB,Serum.TG | 0.012 | 0.273,0.218 | S.LDL.C | 0.059 | 0.003 | 0.1739 | 0.522 |

Table S9: Sensitivity analysis: Top 10 models ranked by the posterior probability and top 10 risk factors ranked by the marginal inclusion probability after model diagnostics (including $n$ = 144 genetic variants for CARDIoGRAMplusC4D and $n$ = 141 for UK Biobank). Causal effects are log odds ratios for coronary artery disease per 1 standard deviation increase in the risk factor.

| $\sigma$ = 0.1 | | | |
|---|---|---|---|
| # | risk factor | *MIP* | $\hat{\theta}_{\text{MACE}}$ |
| 1 | ApoB | 0.638 | 0.193 |
| 2 | S.HDL.TG | 0.365 | 0.073 |
| 3 | LDL.C | 0.253 | 0.05 |
| 4 | IDL.C | 0.165 | 0.029 |
| 5 | IDL.TG | 0.14 | 0.019 |
| 6 | XXL.VLDL.TG | 0.134 | 0.019 |
| 7 | XS.VLDL.TG | 0.127 | 0.018 |
| 8 | Serum.TG | 0.117 | 0.016 |
| 9 | Serum.C | 0.115 | 0.019 |
| 10 | M.HDL.C | 0.11 | -0.013 |

| $\sigma$ = 0.2 | | | |
|---|---|---|---|
| # | risk factor | *MIP* | $\hat{\theta}_{\text{MACE}}$ |
| 1 | ApoB | 0.759 | 0.307 |
| 2 | S.HDL.TG | 0.261 | 0.061 |
| 3 | LDL.C | 0.144 | 0.031 |
| 4 | XXL.VLDL.TG | 0.099 | 0.018 |
| 5 | IDL.C | 0.091 | 0.018 |
| 6 | Serum.C | 0.076 | 0.016 |
| 7 | M.HDL.C | 0.075 | -0.009 |
| 8 | Serum.TG | 0.071 | 0.011 |
| 9 | S.LDL.C | 0.064 | 0.007 |
| 10 | XS.VLDL.TG | 0.064 | 0.008 |

| $\sigma$ = 0.3 | | | |
|---|---|---|---|
| # | risk factor | *MIP* | $\hat{\theta}_{\text{MACE}}$ |
| 1 | ApoB | 0.818 | 0.355 |
| 2 | S.HDL.TG | 0.201 | 0.048 |
| 3 | LDL.C | 0.105 | 0.022 |
| 4 | XXL.VLDL.TG | 0.072 | 0.014 |
| 5 | IDL.C | 0.064 | 0.012 |
| 6 | Serum.C | 0.061 | 0.014 |
| 7 | M.HDL.C | 0.057 | -0.007 |
| 8 | Serum.TG | 0.051 | 0.008 |
| 9 | HDL.C | 0.048 | -0.007 |
| 10 | S.LDL.C | 0.048 | 0.003 |

| $\sigma$ = 0.5 | | | |
|---|---|---|---|
| # | risk factor | *MIP* | $\hat{\theta}_{\text{MACE}}$ |
| 1 | ApoB | 0.868 | 0.392 |
| 2 | S.HDL.TG | 0.136 | 0.033 |
| 3 | LDL.C | 0.075 | 0.015 |
| 4 | XXL.VLDL.TG | 0.047 | 0.01 |
| 5 | Serum.C | 0.045 | 0.011 |
| 6 | IDL.C | 0.042 | 0.008 |
| 7 | S.LDL.C | 0.04 | 0.001 |
| 8 | M.HDL.C | 0.038 | -0.005 |
| 9 | HDL.C | 0.036 | -0.006 |
| 10 | Serum.TG | 0.035 | 0.006 |

| $\sigma$ = 0.7 | | | |
|---|---|---|---|
| # | risk factor | *MIP* | $\hat{\theta}_{\text{MACE}}$ |
| 1 | ApoB | 0.907 | 0.415 |
| 2 | S.HDL.TG | 0.101 | 0.025 |
| 3 | LDL.C | 0.055 | 0.011 |
| 4 | XXL.VLDL.TG | 0.03 | 0.006 |
| 5 | S.LDL.C | 0.029 | 0 |
| 6 | Serum.C | 0.029 | 0.006 |
| 7 | IDL.C | 0.028 | 0.005 |
| 8 | M.HDL.C | 0.026 | -0.003 |
| 9 | Serum.TG | 0.023 | 0.004 |
| 10 | HDL.C | 0.023 | -0.003 |

Table S10: Parameter check for the prior variance $\sigma^2$, ranging from $\sigma = 0.1$ to $\sigma = 0.7$. The main analysis used $\sigma = 0.5$. Abbreviations: *MIP*=marginal inclusion probability, MACE=model-averaged causal effect.

| $p$ = 0.01 | | | |
|---|---|---|---|
| # | risk factor | *MIP* | $\hat{\theta}_{\text{MACE}}$ |
| 1 | ApoB | 0.979 | 0.454 |
| 2 | S.HDL.TG | 0.015 | 0.004 |
| 3 | LDL.C | 0.009 | 0.002 |
| 4 | S.VLDL.C | 0.007 | 0.002 |
| 5 | S.LDL.C | 0.004 | 0 |
| 6 | Serum.C | 0.004 | 0.001 |
| 7 | XS.VLDL.TG | 0.004 | 0.001 |
| 8 | IDL.C | 0.004 | 0.001 |
| 9 | M.HDL.C | 0.004 | 0 |
| 10 | XXL.VLDL.TG | 0.004 | 0.001 |

| $p$ = 0.05 | | | |
|---|---|---|---|
| # | risk factor | *MIP* | $\hat{\theta}_{\text{MACE}}$ |
| 1 | ApoB | 0.929 | 0.426 |
| 2 | S.HDL.TG | 0.071 | 0.017 |
| 3 | LDL.C | 0.039 | 0.008 |
| 4 | Serum.C | 0.022 | 0.005 |
| 5 | S.LDL.C | 0.02 | 0.001 |
| 6 | XXL.VLDL.TG | 0.02 | 0.004 |
| 7 | IDL.C | 0.02 | 0.004 |
| 8 | M.HDL.C | 0.019 | -0.002 |
| 9 | HDL.C | 0.017 | -0.003 |
| 10 | S.VLDL.C | 0.016 | 0.001 |

| $p$ = 0.1 | | | |
|---|---|---|---|
| # | risk factor | *MIP* | $\hat{\theta}_{\text{MACE}}$ |
| 1 | ApoB | 0.868 | 0.392 |
| 2 | S.HDL.TG | 0.136 | 0.033 |
| 3 | LDL.C | 0.075 | 0.015 |
| 4 | XXL.VLDL.TG | 0.047 | 0.01 |
| 5 | Serum.C | 0.045 | 0.011 |
| 6 | IDL.C | 0.042 | 0.008 |
| 7 | S.LDL.C | 0.04 | 0.001 |
| 8 | M.HDL.C | 0.038 | -0.005 |
| 9 | HDL.C | 0.036 | -0.006 |
| 10 | Serum.TG | 0.035 | 0.006 |

| $p$ = 0.2 | | | |
|---|---|---|---|
| # | risk factor | *MIP* | $\hat{\theta}_{\text{MACE}}$ |
| 1 | ApoB | 0.791 | 0.347 |
| 2 | S.HDL.TG | 0.238 | 0.059 |
| 3 | LDL.C | 0.127 | 0.025 |
| 4 | XXL.VLDL.TG | 0.099 | 0.022 |
| 5 | Serum.C | 0.083 | 0.019 |
| 6 | IDL.C | 0.076 | 0.013 |
| 7 | Serum.TG | 0.071 | 0.014 |
| 8 | S.LDL.C | 0.07 | 0.001 |
| 9 | M.HDL.C | 0.068 | -0.008 |
| 10 | HDL.C | 0.068 | -0.01 |

| $p$ = 0.3 | | | |
|---|---|---|---|
| # | risk factor | *MIP* | $\hat{\theta}_{\text{MACE}}$ |
| 1 | ApoB | 0.744 | 0.318 |
| 2 | S.HDL.TG | 0.32 | 0.081 |
| 3 | XXL.VLDL.TG | 0.18 | 0.046 |
| 4 | LDL.C | 0.164 | 0.032 |
| 5 | Serum.TG | 0.117 | 0.025 |
| 6 | Serum.C | 0.11 | 0.023 |
| 7 | IDL.C | 0.106 | 0.018 |
| 8 | S.VLDL.C | 0.103 | -0.014 |
| 9 | M.HDL.C | 0.092 | -0.011 |
| 10 | S.LDL.C | 0.092 | 0.001 |

Table S11: Parameter check for the prior probability $p$, ranging from $p = 0.01$ to $p = 0.3$. This reflects 0.3 to 9 expected causal risk factors. The main analysis used $p = 0.1$ reflecting an a priori expected number of 3 causal risk factors. Abbreviations: *MIP*=marginal inclusion probability, MACE=model-averaged causal effect.

### Supplementary Figures

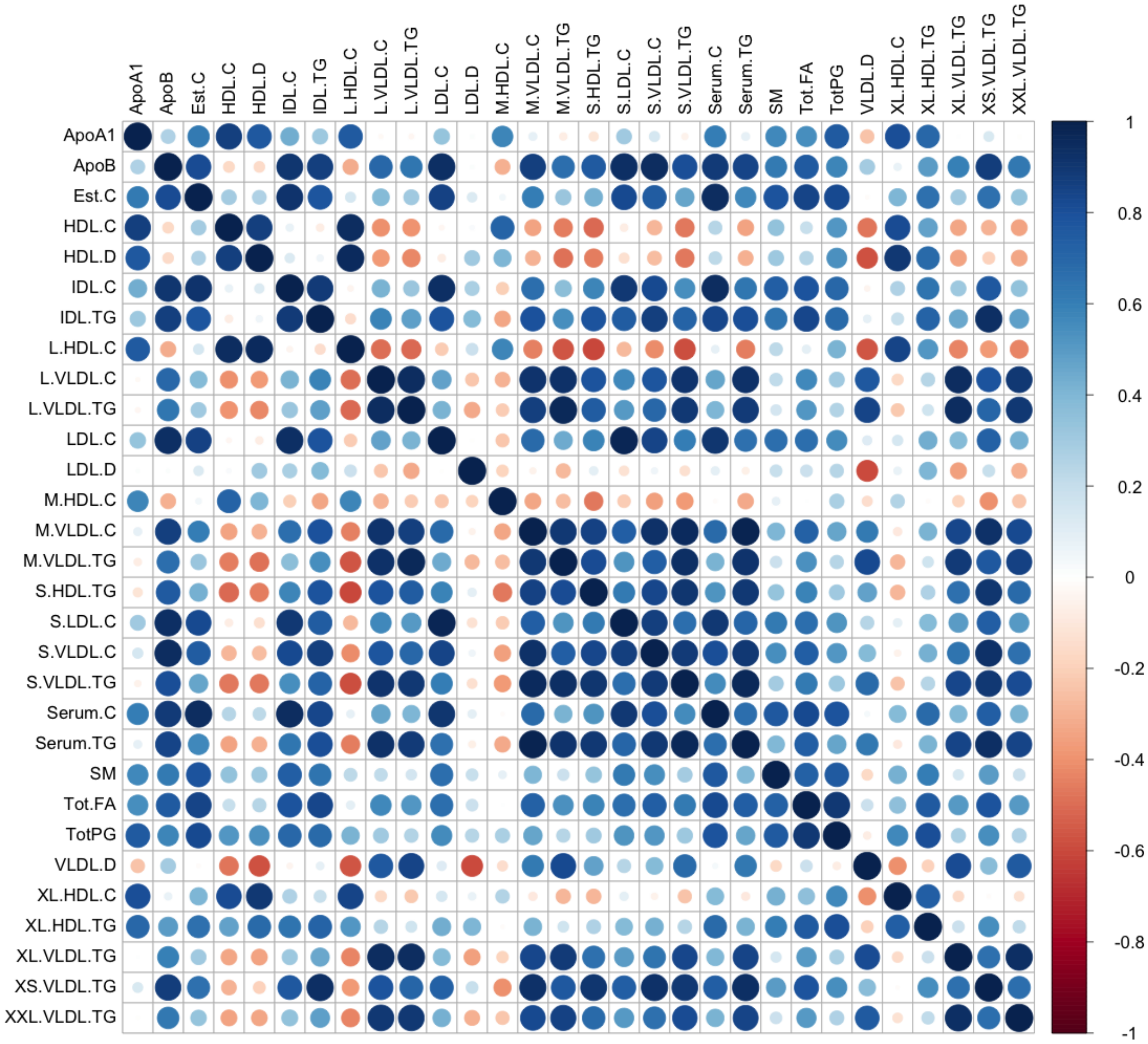


Figure S1: Genetic correlation between lipoprotein measures and metabolites based on the $n$ = 148 lipid-associated genetic variants as used in the main analysis. Colorcode indicates correlation strength (darkblue=strong positive correlation to darkred=strong negative correlation). The size of the square is proportional to the absolute correlation.

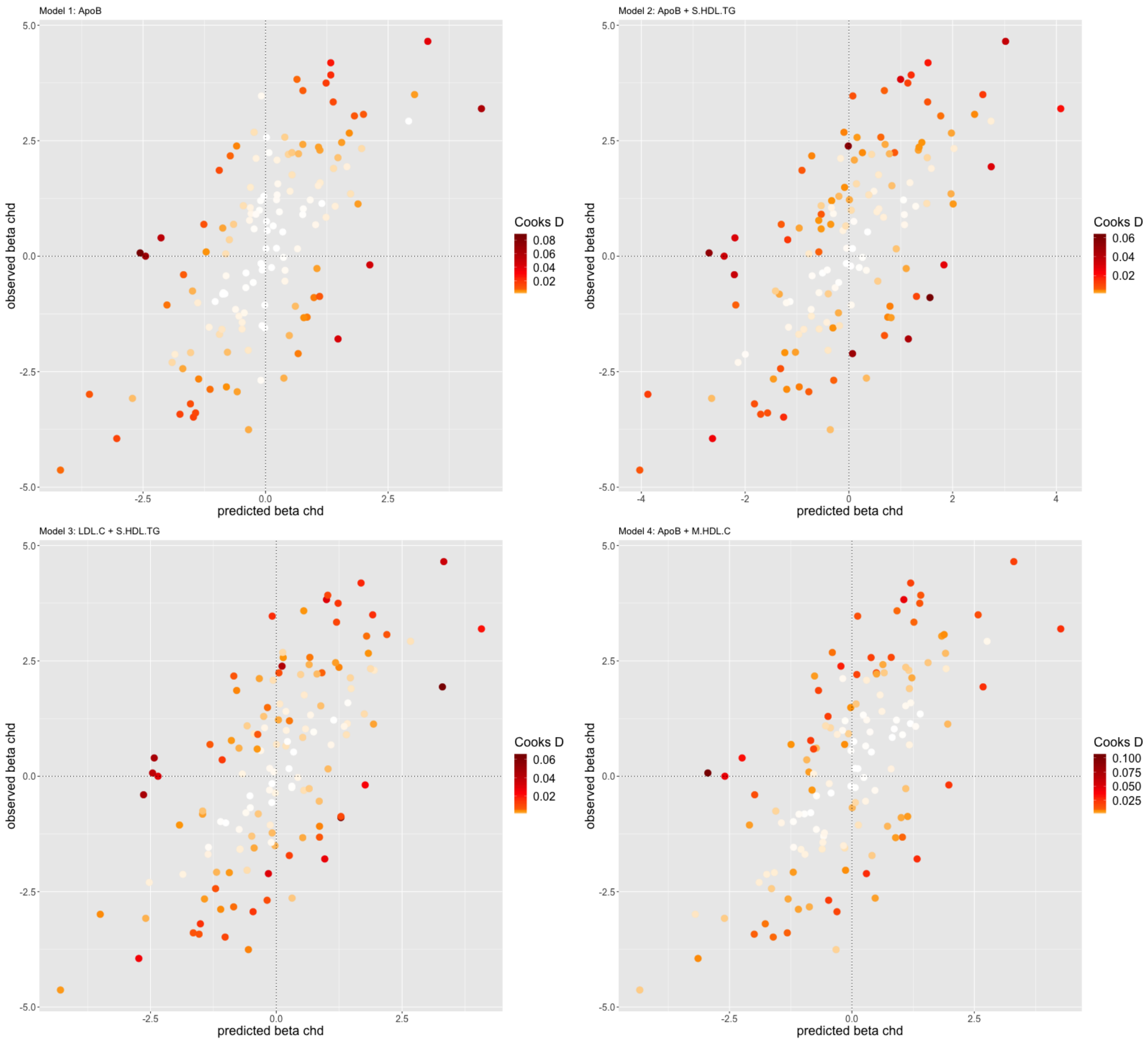


Figure S2: Diagnostic plots with Cooks distance: Estimates of genetic associations with the outcome against predicted genetic associations with the outcome from the primary analysis based on $n$ = 138 genetic variants after exclusion of outliers. Here we show the diagnostics for all four top models with posterior probability > 0.02 as given in Main Table 1.Colour code of points indicates influence, as measured by the variant's Cook's distance.

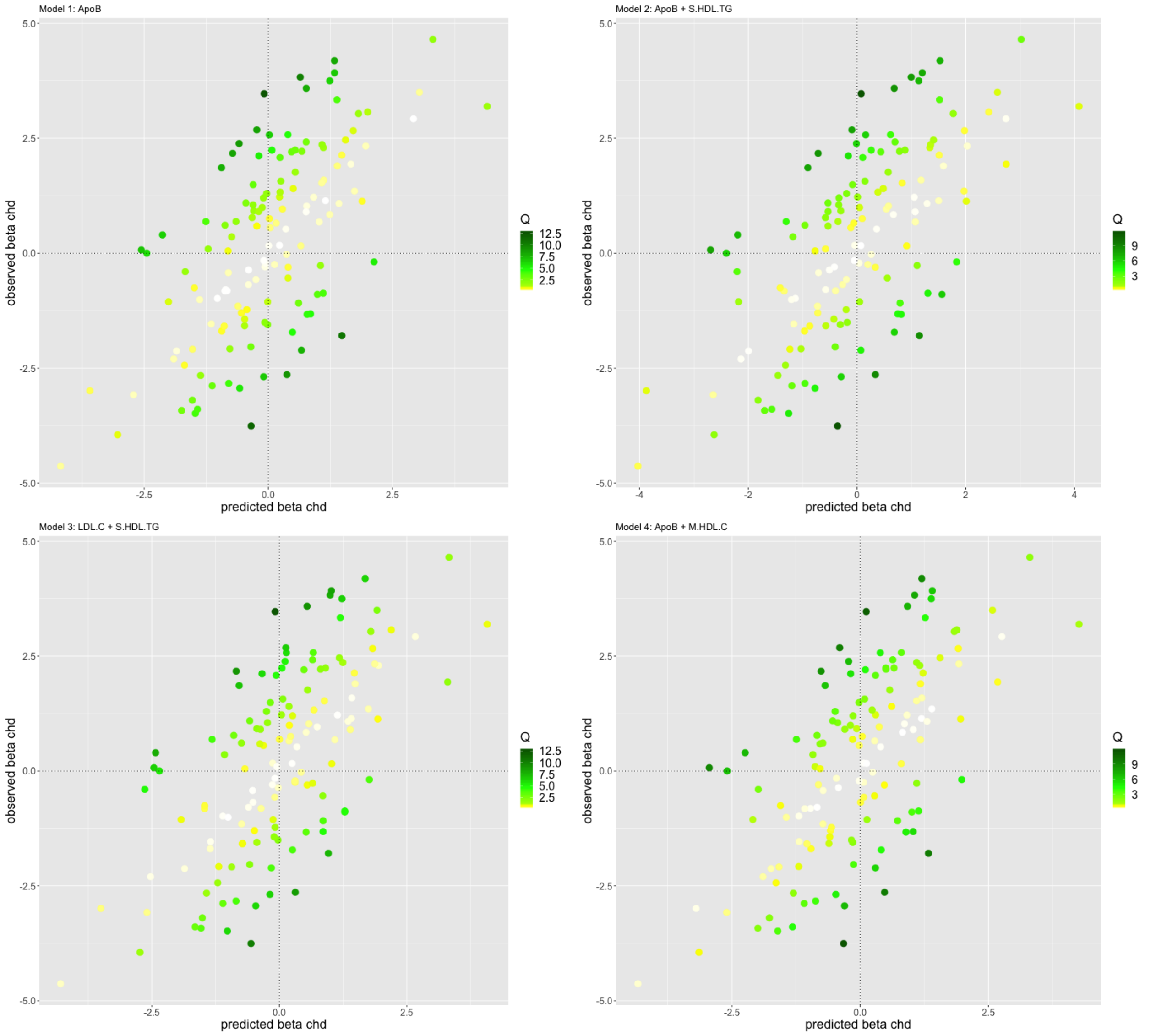


Figure S3: Diagnostic plots with $q$-statistic: Estimates of genetic associations with the outcome against predicted genetic associations with the outcome from the primary analysis based on $n = 138$ genetic variants after exclusion of outliers. Here we show the diagnostics for all four top models with posterior probability $> 0.02$ as given in Main Table 1. Colour code of points indicates heterogeneity, as measured by the variant's $q$-statistic.

### Supplementary Methods

#### Mendelian randomization using summarized data

A genetic variant can be used to make causal inferences about the effect of a risk factor on an outcome if it satisfies the three instrumental variable assumptions:

IV1 The variant is associated with the risk factor;

IV2 The variant is not confounded in its associations with the outcome;

IV3 The variant does not influence the outcome directly, only potentially indirectly via its association with the risk factor.

These assumptions imply that a genetic variant behaves analogously to random assignment to a treatment group in a randomized controlled trial, in that it divides the population into subgroups that differ only with respect to their average level of the risk factor [1]. Any difference in the outcome between these groups implies a causal effect of the risk factor on the outcome, analogous to an intention-to-treat effect in a randomized trial [2].

We consider an extension of the Mendelian randomization paradigm known as multivariable Mendelian randomization, in which genetic variants are allowed to influence multiple risk factors, provided that any causal pathway from the genetic variants to the outcome passes via one or more of the measured risk factors [3]. The assumptions for genetic variants to be valid instruments in multivariable Mendelian randomization are:

MV-IV1 Each variant is associated with at least one of the risk factors;

MV-IV2 Variants are not confounded in their associations with the outcome;

MV-IV3 Variants are not associated with the outcome conditional on the risk factors and confounders.

In turn, the assumptions for a risk factor to be included in a multivariable Mendelian randomization model are:

RF1 No risk factor can be linearly explained by any other included risk factor or a combination of multiple risk factors.

RF2 Each risk factor is associated with at least one of the genetic variants.

Assumption RF1 is needed to distinguish between correlated risk factors [4]. RF2 ensures that each risk factor is adequately predicted by the genetic variants selected as instrumental variables in the analysis.

For a particular set of risk factors, causal effects are estimated by weighted linear regression of the genetic associations with the outcome on the genetic associations with the risk factors

$$\beta_Y = \theta_1 \beta_{X1} + \theta_2 \beta_{X2} + \ldots + \theta_d \beta_{Xd} + \varepsilon, \quad \varepsilon \sim N(0, \text{diag}(\text{se}(\beta_Y)^2)),$$

where $\beta_Y$ is the vector of genetic associations with the outcome of length $n$, with $n$ the number of genetic variants used as instrumental variables, $\text{se}(\beta_Y)$ is the vector of standard errors of these associations of length $n$ and diag the diagonal operator. $\beta_{X1}, \beta_{X2}, \ldots, \beta_{Xd}$ are the genetic associations with the $d$ risk factors, and $\theta_1, \theta_2, \ldots, \theta_d$ are the causal effects of the $d$ risk factors on the outcome. If there are causal relationships between the risk factors, then these parameters represent the direct effects of the risk factors, i.e. the effect of changing the target risk factor keeping all other risk factors constant [4, 5].

#### Variable selection and Bayesian model averaging

The model averaging approach is implemented by considering different sets of risk factors in turn [6]. For each risk factor set, MR-BMA fits the relevant multivariable Mendelian randomization model and assigns a score to the set of risk factors considered that captures the posterior probability that this particular model represents the true causal risk factors for the outcome given the observed genetic association data [6]. As prior parameters MR-BMA requires to set an a priori probability for a risk factor to be causal, which is set to 0.1 reflecting an a priori epectation of three causal risk factors. Additionally, the prior variance is set to 0.25. Sensitivity analysis with respect to the prior parameters is important and we can show that ranking is not impacted by the choice of the prior. Results for a wide range of prior specifications are given in Supplementary Table S10 (prior variance) and Supplementary Table S11 (prior probability).

When considering many candidate risk factors, the model space (including all possible combinations of risk factors) may be prohibitively large to consider all possible combinations of risk factors. To alleviate this we have implemented a stochastic search algorithm [7] to explore the relevant model space (all models with a non-negligible posterior probability) in an efficient way.

When the number of risk factors considered is large, the evidence for each particular model may be small. Hence, we average over the models visited and for each risk factor compute its marginal inclusion probability, which is the sum of the posterior probabilities for all models visited that include this particular risk factor. Further, we provide the model-averaged causal effect estimate, representing the average causal effect estimate for the given risk factor across models in which it is included. As is common for variable-selection methods, this is a conservative estimates of the true causal effect and underestimates its magnitude, but may be used for the interpretation of effect direction and for comparison among the risk factors.

#### Resampling to compute empirical $p-$values

Empirical $p$-values for the marginal inclusion probability of each risk factor are obtained using a permutation procedure, where the risk factor association data are held constant and the outcome associations of the genetic variants are randomly perturbed [8]. The empirical $p$-value for risk factor $j$ quantifies how extreme the actual observed marginal inclusion probability is with respect to all permuted marginal inclusion probabilities for that particular risk factor. Formally, the empirical $p$-value is computed by the rank ($r_j$) of the actual observed marginal inclusion probability for risk factor $j$ among all permuted marginal inclusion probabilities for risk factor $j$ over the total number of permutations ($n_{perm}$ = 1,000). Following [9] we add one to the computation to obtain the probability that under the null hypothesis the observed marginal inclusion probability has the observed or a higher rank

$$p_j = (r_j + 1)/(n_{perm} + 1).$$

Multiple testing adjustment is done using the Benjamini and Hochberg false discovery rate (FDR) procedure [10].

#### Model diagnostics

Two approaches are considered for model diagnostics. Firstly, to identify influential variants for each visited model with a model posterior probability larger than 0.02, we calculated Cook's distance for each genetic variant [11] and excluded all variants that have in any selected model a Cook's distance which exceeds the median of a central $F$-distribution with $d$ and $n-d$ degrees of freedom, where $d$ is the number of risk factors and $n$ the number of genetic variants used as instrumental variables.

Secondly, to identify outlying variants, we consider for each visited model with a model posterior probability larger than 0.02 a version of Cochran's Q statistic used to detect heterogeneity in meta-analysis [12]

$$Q = \sum_{i=1}^{n} q_i = \sum_{i=1}^{n} \mathrm{se}(\beta_{Y_i})^{-2}(\beta_{Y_i} - \hat{\beta}_{Y_i})^2,$$

where $i$ indexes the genetic variants and $\hat{\beta}_{Y_i}$ is the predicted value of the genetic association with the outcome $\beta_{Y_i}$ based on the relevant multivariable Mendelian randomization model. A genetic variant with a high value of $q_i$ (compared to the $0.05/n$th upper tail of a $\chi^2$ distribution with one degree of freedom representing Bonferroni multiple testing adjustment by the number of variants included) in any of the models visited (with a model posterior probability larger than 0.02) was considered to be an outlying variant.

We then repeated the analyses excluding such variants. The reason for excluding outliers and influential variants is that a single genetic variant can have a strong impact on the models visited and subsequently on variable selection. However, in this case for both main and sensitivity analyses, excluding these variants did not change the headline results.
